# Potential Therapeutic Options for Ethanol-Producing Ethanol-Resistant Gut Microbes associated with Liver Diseases

**DOI:** 10.1101/2025.11.13.25340061

**Authors:** Anissa Idrissa Abdoulaye, Babacar Mbaye, Reham Magdy Wasfy, Louis Carmarans, Mamadou Beye, Claudia Andrieu, Sofiane Bakour, Aïcha Hamieh, Nicholas Armstrong, Patrick Borentain, Stéphane Ranque, Jean-Marc Rolain, Gregory Dubourg, Jean-Christophe Lagier, Maryam Tidjani Alou, René Gerolami, Matthieu Million

## Abstract

**Background:** Endogenous ethanol (EtOH) production is an emerging pathophysiological mechanism involved in metabolic dysfunction-associated steatohepatitis (MASH). Therefore, characterizing EtOH-producing species enriched in patients with liver disease could help identify putative pathobionts and potential therapeutic treatments.

**Methods:** We investigated EtOH production and tolerance, antimicrobial susceptibility and antimicrobial resistance gene(s) in 33 strains enriched in MASH, alcoholic hepatitis (AH) and HBV patients. An antimicrobial was considered a potential therapeutic option when the ratio of the fecal concentration/minimum inhibitory concentration (FC/MIC) was > 10 (1 log_10_).

**Results:** 92% of species enriched in patients with liver diseases produced detectable amounts of ethanol with a strong association between EtOH tolerance and production (p < 0.05). *Candida albicans*, *Nakaseomyces glabratus* and *Pichia kudriavzevii* produced the highest concentrations of EtOH (1.8 to 3.3 g/L). *Enterocloster*, a strictly anaerobic bacterial genus, was the bacterial genus with the highest EtOH production (0.8 to 1.6 g/L) in 5 g/L of glucose. Poorly absorbed drugs, amphotericin B, rifaximin and vancomycin, together constituted potential therapeutic options for all tested strains.

**Conclusions:** In addition to yeasts, *Lactobacillaceae* and *Klebsiella*, strains of *Enterocloster* genus isolated from patients with liver diseases produced significant amount of EtOH. Most gut microbial species associated with liver diseases produce and tolerate ethanol, suggesting a possible vicious microbial-ethanol cycle in such context. Finally, non-absorbed antimicrobials (rifaximin, vancomycin and amphotericin B) already used for gut microbiota-targeted therapies may open new avenues in the design of precision medicine.

## Introduction

Liver diseases are a major public health problem. A study estimated that in 2023, over 2 million deaths or 4% of the global mortality rate, are caused by cirrhosis, viral hepatitis and liver cancer annually (1). The main causes of cirrhosis are chronic infection with hepatitis B virus (HBV) or hepatitis C virus (HCV), alcoholic hepatitis (AH), and metabolic dysfunction-associated steatohepatitis (MASH) (2). The emergence of metabolic hepatic diseases, including metabolic dysfunction-associated fatty liver disease (MAFLD) and its severe form, MASH, is growing worldwide. Indeed, in 2019, the estimated prevalence of MAFLD was 1.6 billion worldwide (3). It increased from 25% between 1990 and 2006 to 38% between 2016 and 2019 (4). The global prevalence of MASH is estimated to be 5%, with the highest prevalence in Latin America (44%) (4).

Alterations in the gut microbiota have been observed in viral hepatitis (5,6), AH (7), MAFLD and MASH (8). Moreover, one feature of MASH is endogenous ethanol production (9), which was initially demonstrated by Yuan et al. with *Klebsiella pneumoniae* strains isolated from patients with auto-brewing syndrome (ABS) and MASH (10). Similarly, Meijnikman et al. reported a positive correlation between the abundance of *Lactobacillaceae* and endogenous ethanol production in MAFLD patients (11). Interestingly, the causal role of these microbes in endogenous ethanol production was confirmed through *in vitro* and *in vivo* experimental studies (10,12). Moreover, Casanas et al. recently treated a patient with metabolic dysfunction-associated steatosis liver disease (MASLD) and ABS using fecal microbiota transplantation (FMT) (13).

While prior research has established aerobic species-mediated endogenous ethanol production in MASH and ABS (10,12), our culturomics approach applied to the gut microbiota from MASH and viral hepatitis patients also highlighted enriched anaerobic bacterial species. These studies revealed enrichment of yeasts in the intestinal microbiota of MASH patients, specifically *Candida albicans*, *Nakaseomyces glabratus* and *Pichia kudriavzevii* (14). Similarly, enriched bacterial species included *Limosilactobacillus fermentum*, *Mediterraneibacter gnavus*, *Streptococcus mutans* and *Enterocloster bolteae* in the intestinal microbiota of MASH and HBV patients (15,16), as well as enrichment of *Thomasclavelia ramosa* in the intestinal microbiota of patients with alcohol-related hepatocellular carcinoma (17).

As the etiology of MASH and ABS is related to endogenous ethanol production by gut microbial taxa, microbiota-targeted therapies might be relevant for these pathologies (9,18). The identification and characterization of potential microbial biomarkers represent crucial steps for that purpose (19). In this study, we investigated ethanol production, ethanol tolerance, antibiotic susceptibility and antibiotic resistance gene expression in thirty-three ethanol-producing strains that were enriched and isolated from patients afflicted with MASH, AH and HBV.

## Materials and methods

### Ethical approval

All the strains used in this study were isolated in the HEPATGUT project, which was approved by the ethics and personal protection committees (CPP: 21.04391.000046 - 21075). In accordance with the Declaration of Helsinki (World Medical Association, 2013), written informed consent was obtained from each participant.

### Studied strains

Thirty-three strains, including bacteria and yeasts, obtained from culturomics case–control studies on MASH, HVB and AH patients conducted in our laboratory and significantly enriched in these pathologies were included in this study (14–16). Strains of species enriched in patients and those enriched in controls were studied regardless of the sample of origin (Table 1). Hence, we included the following species enriched in the intestinal microbiota of MASH patients: *Bacteroides thetaiotaomicron*, *C. albicans, E. bolteae*, *Enterocloster clostridioformis*, *K. pneumoniae*, *Klebsiella michiganensis*, *L. fermentum*, *M. gnavus*, *N. glabratus*, *Peptoniphilus grossensis, P. kudriavzevii* and *T. ramosa* (14,15,20). Strains of *T. ramosa* enriched in the gut microbiota of patients with AH and chronic HBV infection were also studied (17). In addition, species significantly depleted in the intestinal microbiota of MASH patients compared to healthy individuals were included, specifically *Alistipes shahii* and *Bacteroides uniformis* (15). In fact, *A. shahii* is known to have beneficial effects on various pathologies, including liver fibrosis (21). Moreover, *B. uniformis* is a known gut commensal that has been shown to alleviate MASH (22).

**Table 1.**
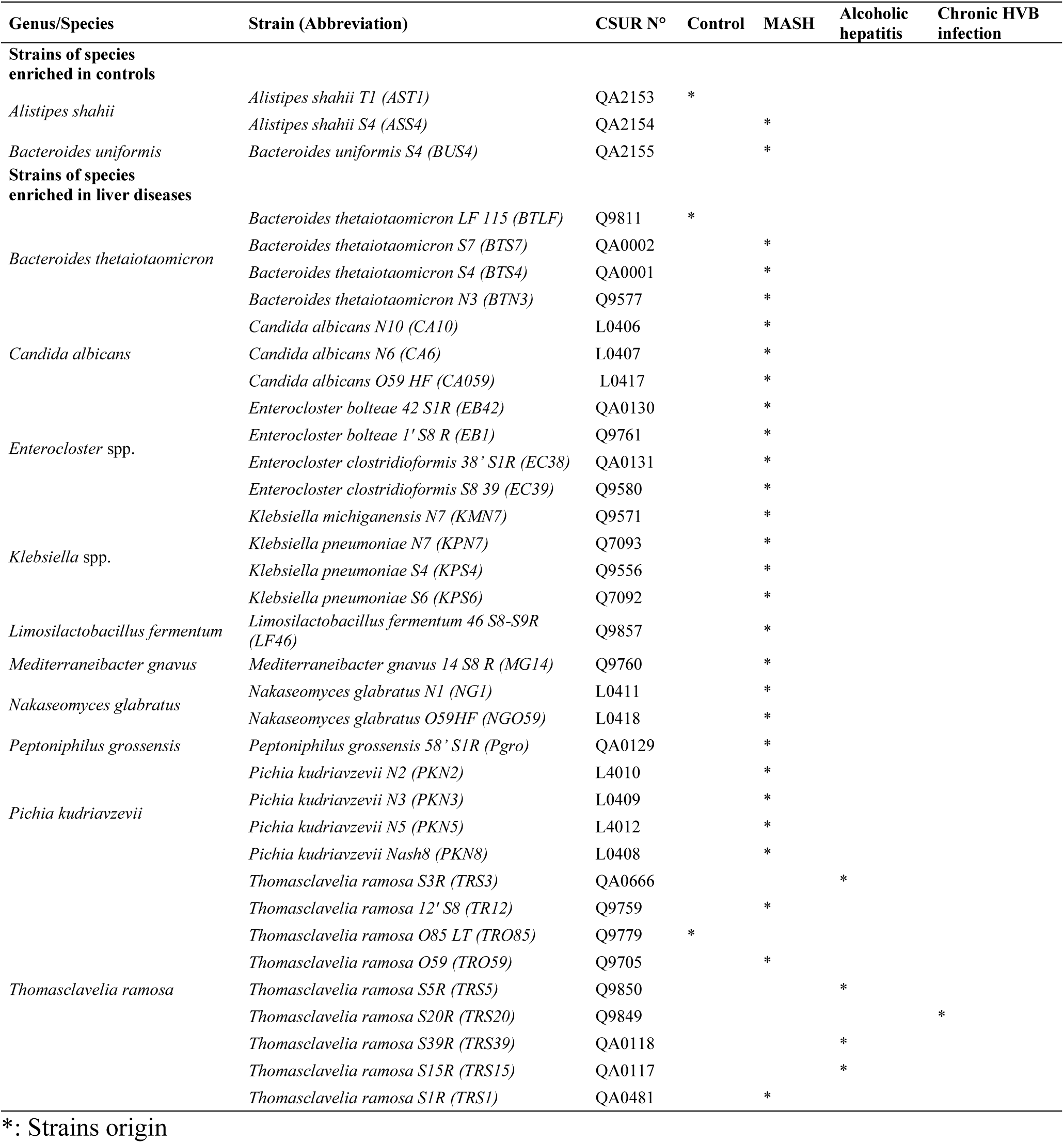
Strains of interest.

### *In vitro* ethanol production assay

For each species, ethanol production was quantified in triplicate in the initial isolation medium. Hence, bacteria were grown in Columbia broth base enriched with 5% defibrinated sheep blood (COS) at 37°C for 24 hours for aerobic bacteria and 48 hours for anaerobic bacteria. Yeasts were grown in Sabouraud broth (Oxoid, Basingstoke, UK) at 30°C for 24 hours. One milliliter of each culture and of a negative control (sterile culture medium) were subsequently transferred to 20 mL glass Headspace vials (Supplementary materials and methods).

Considering the difference in glucose concentration between the COS (5 g/L) used for bacteria and Sabouraud broth (20 g/L) used for yeast, the highest concentration of ethanol-producing bacterial strains was also tested using modified COS broth (COS with 20 g/L glucose).

The ethanol concentration was determined using a headspace gas chromatography–mass spectrometry (HS–GC–MS) system (Perkin Elmer, Villebon sur Yvette, France) in the Swafer D7 setup, which included an HS110 headspace injector, a Clarus 690 gas chromatograph and an SQ8T mass spectrometer, as described by Sissoko et al. (23).

### Ethanol tolerance test

The ethanol tolerance of bacteria was assessed in COS broth containing 0%, 5% and 10% ethanol, whereas that of yeast was assessed in Sabouraud broth (Oxoid) supplemented with 0%, 5% and 10% ethanol (Supplementary materials and methods).

### Statistics

To compare quantitative variables, Mann–Whitney or Student’s t tests were applied on the basis of their distribution. All tests were two-tailed. A p value < 0.05 was used to determine significance. All statistical analyses were performed using GraphPad Prism version 10.4.1 (GraphPad Software, Boston, Massachusetts USA, www.graphpad.com).

### Antifungal and antibiotic susceptibility tests

Antifungal susceptibility testing to amphotericin B, caspofungin, flucytosine, micafungin and voriconazole was conducted using the VITEK 2 automated system (bioMérieux, Marcy l’Etoile, France) and VITEK 2 AST-YS08 antifungal cards (bioMérieux, Durham, USA). Susceptibility to fluconazole was assessed using E-test strips (bioMerieux, Marcy l’Etoile, France) (Supplementary materials and methods).

The susceptibility to 25 antibiotics was assessed with the disc diffusion method using E-test strips (bioMerieux) (Supplementary materials and methods). Considering the widespread use of rifaximin in the treatment of patients with liver disease, we also assessed their susceptibility to rifaximin. The minimum inhibitory concentration (MIC) of rifaximin for each strain was assessed using the agar dilution method, according to the recommendations of the Clinical Laboratory Standards Committee (CLSI), as there were no E-test strips commercially available for this molecule.

### Potential efficacy of rifaximin, vancomycin and amphotericin B *in vivo*

To determine the potential efficacy of antimicrobials on the species mentioned above, we selected the least absorbable molecules (amphotericin B, rifaximin and vancomycin) and compared their fecal concentrations (FC) reported in the literature (24–26) to the MICs obtained in this study (Supplementary materials and methods). This ratio (FC/MIC) aim to determine the potential efficacy of these drugs in liver diseases associated to endogenous ethanol production, as our target is the gut microbiota. We established an FC/MIC threshold equal to 10, which indicates an efficacy of antimicrobials on species targeted in gut microbiota.

### Antimicrobial(s) resistance gene(s)

After bacterial genome sequencing and assembly (Supplementary materials and methods), the presence of antimicrobial resistance gene(s) was assessed by the command abritAMR with AMRFinder Plus via Galaxy Australia (https://usegalaxy.org.au/).

Additionally, for strains presenting a high resistance to rifampin and rifaximin (> 256 µg/mL), the presence of mutation(s) in the rifampin resistance-determining region (RRDR) in the β subunit of bacterial RNA polymerase (rpoB) which is the main reported cause of resistance to rifampicin (27) was assessed after genomes annotation with PROKKA via Galaxy Australia (https://usegalaxy.org.au/, last accessed on October 16^th^, 2025) and rpoB alignment in MEGA 12 (version 12.0.14).

## Results

### High ethanol production by microbial species associated with liver diseases

The production of ethanol by strains of species enriched in the intestinal microbiota of the controls (*A. shahii* and *B. uniformis*) was low, always under 0.03 g/L (Table S1), compared with that of species enriched in the intestinal microbiota of liver disease patients (n= number of strains, median= median of ethanol dosed in g/L [interquartile range], n= 33, 0.58 [0.21–1.86] vs. n=3, 0.026 [0.025–0.028], 22.3-fold, two-tailed Mann–Whitney test, p <0.0001, Figure S1). Indeed, ethanol production was 22-fold greater in species enriched in the intestinal microbiota of patients with liver diseases than in those enriched in the intestinal microbiota of the controls. A total of 92% (11/12) of the species associated with liver diseases produced detectable levels of ethanol, whereas none of the strains enriched in the controls (0/3) produced detectable levels of ethanol. *B. thetaiotaomicron* was the only species enriched in the microbiota of patients with liver diseases that did not produce any detectable amount of ethanol.

The analysis of the amount of ethanol produced by each strain revealed that compared to the bacterial strains, the yeast strains produced 11 times more ethanol (n=9, 2.70 [2.11–2.98] vs. n=27, 0.24 [0.03–0.61], 11.25-fold, two-tailed Mann–Whitney test; p <0.0001) (Figure S2). The two strains of *N. glabratus* produced the greatest amount of ethanol (3.29 g/L and 3.06 g/L), followed by the *P. kudriavzevii* strains (2.89 g/L–2.48 g/L) and *C. albicans* strains (2.34 g/L–1.84 g/L) (Figure 1.A).

**Figure 1.**
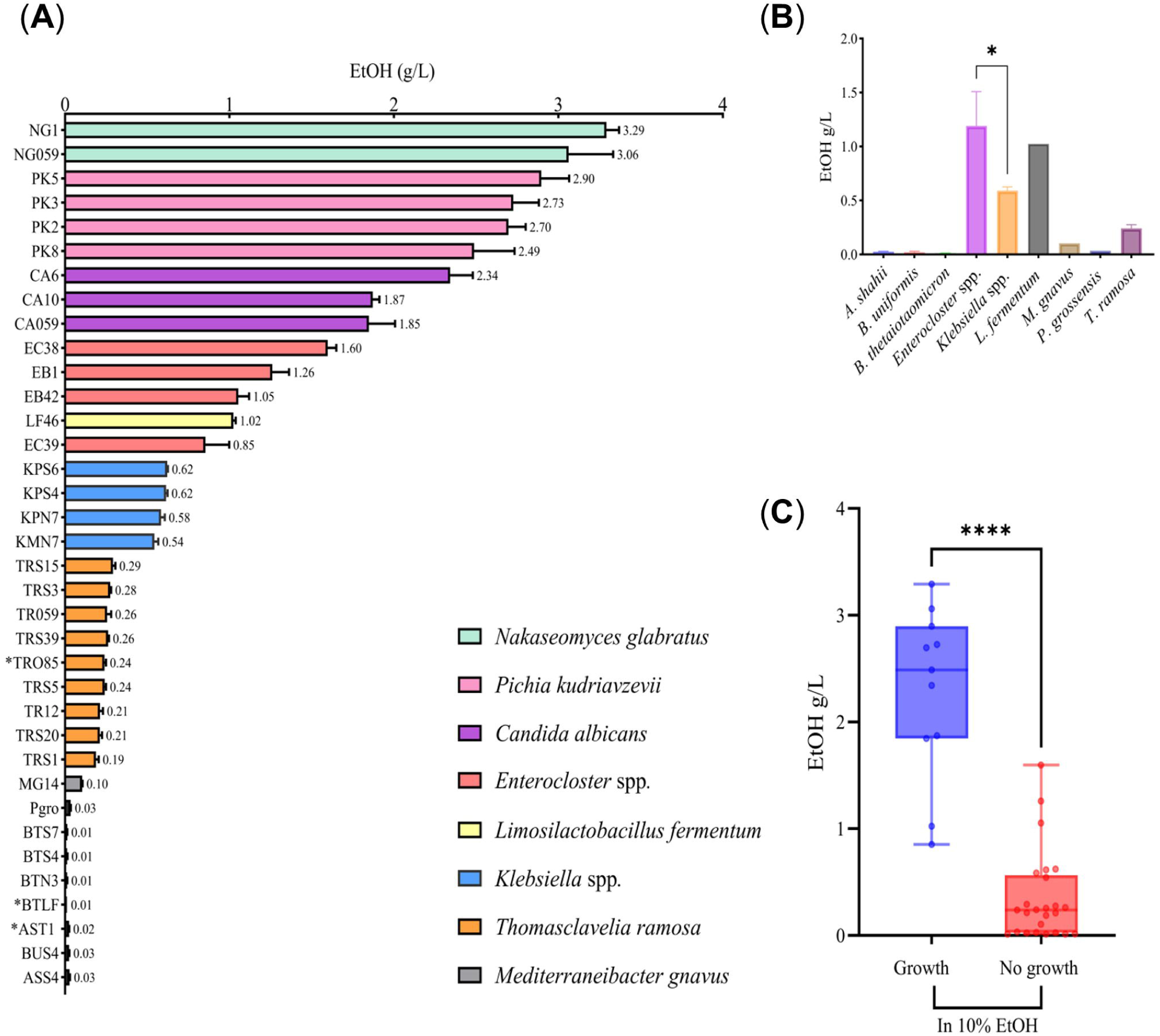
Ethanol production by yeast and bacteria dosed by GC–MS and their ethanol tolerance. (A) NG: *N. glabratus*, CA: *C. albicans*, PK: *P. kudriavzevii*, KM: *K. michiganensis*, KP: *K. pneumoniae*, BU: *B. uniformis*, AS: *A. shahii*, BT: *B. thetaiotaimicron*, MG: *M. gnavus*, LF: *L. fermentum*, Pgro: *P. grossensis*, EB: *E. bolteae*, EC: *E. clostridioformis*, TR: *T. ramosa*, EtOH: Ethanol quantity dosed, *: Strains isolated from controls. (B) Ethanol production by species (EtOH: Ethanol produced dosed). Level of significance: *: p = 0.0286 (two-tailed Mann–Whitney test). (C) Comparison of strains that grew in 10% ethanol medium with those that did not grow in terms of ethanol production (EtOH: Ethanol quantity dosed). Level of significance: ****: p<0.0001 (two-tailed Mann–Whitney test).

The two *E. bolteae* strains produced ethanol concentrations greater than 1 g/L (1.26 g/L and 1.05 g/L), as did the two *E. clostridioformis* strains (1.59 g/L and 0.85 g/L). *L. fermentum* produced 1.02 g/L ethanol. All the strains of *Klebsiella* spp. produced ethanol at concentrations above 0.5 g/L (0.54–0.62 g/L). *T. ramosa* strains produced ethanol concentrations ranging from 0.18 g/L to 0.29 g/L. *M. gnavus* (MG14) and *P. grossensis* (Pgro) produced 0.10 g/L and 0.03 g/L ethanol, respectively. *B. thetaiotaomicron* strains produced the lowest concentration (0.01 g/L) (Figure 1.A). Notably, strains from the same species produced similar amounts of ethanol, suggesting an overall species-dependent effect (Figure 1A). *Enterocloster* spp. produced twice as much ethanol as *Klebsiella* spp. did (Figure 1.B) (n=4, 1.16 [0.90–1.51]; n=4, 0.59 [0.55–0.62]; 1.97-fold, two-tailed Mann–Whitney test; p=0.0286). The ethanol concentration measured in sterile media was consistently under 0.006 g/L (Table S2).

As Sabouraud broth, which is used for yeast culture, contains 20 g/L glucose, we also assessed the ethanol production of the highest bacterial ethanol producers of each species in modified COS broth with 20 g/L of glucose, namely, *B. thetaiotaomicron* (BTN3), *E. clostridioformis* (EC38), *K. pneumoniae* (KPS6), *L. fermentum* (LF46) and *T. ramosa* (TRS5, Figure S3.A). The increased glucose concentration resulted in an increased ethanol production in some species ((*L. fermentum* (LF46) and *K. pneumoniae* (KPS6)), whereas it resulted in a decreased ethanol production in others ((*E. clostridioformis* (EC38) and *T. ramosa* (TRS5)) (Figure S3.A). A comparison of ethanol production between bacterial and yeast strains in the presence of the same amount of glucose in the medium (20 g/L) (Figure S3.B) revealed that compared with bacteria, yeast still produced significantly more ethanol (n=3, 2.8 [2.40–3.25] vs. n=5, 0.58 [0.12–0.96], 4.8-fold, two-tailed Mann–Whitney test; p <0.0001). At 20 g/L, the same gradient of ethanol production per species was maintained, except for *E. clostridioformis* (EC38), which produced less ethanol than *K. pneumoniae* (KPS6) did (Figure S3.B).

### High ethanol production is associated with high ethanol tolerance

All yeast strains grew in 10% ethanol, as did two bacterial strains, *L. fermentum* (LF46) and *E. clostridioformis* (EC39) (Table S3). All bacterial strains grew at a concentration of 5% ethanol, except *E. bolteae* (EB42) and *A. shahii* (ASS4) (Table S3). None of the species enriched in the intestinal microbiota of the controls grew in the presence of 10% ethanol (Table S3). The strains that grew in 10% ethanol produced significantly more ethanol (p < 0.0001) than those that did not (Figure 1.C). Those that were tolerant to 10% ethanol produced ten times more ethanol than those that were not (n = strains, m= median of ethanol dose in g/L [inter quartile range], n=11, 2.49 [1.85–2.90] vs. n=25, 0.24 [0.03–0.56], 10.4-fold, two-tailed Mann–Whitney test, p <0.0001) (Figure 1.C).

### High potential activity of amphotericin B, rifaximin and vancomycin *in vivo*

The detailed antibiotic and antifungal susceptibility results are reported in Table S4 and Table S5. Antibiotic susceptibility is usually determined according to standards recommended by the EUCAST in Europe, and the MICs are determined according to concentrations of the molecule of interest in the blood, which is not relevant for our specific target, which is the gut microbiota. Therefore, it is important to assess the potential efficacy of molecules of interest in the gut as we target gut commensals. Hence, based on four characteristics for each antibiotic (no absorbability by the intestinal barrier, a low MIC, a fecal concentration available in the literature dosed in humans and availability of an oral galenic form), we selected antimicrobials that could have high activity *in vivo* (Table S6).

In fact, EUCAST recommendations are generally available only for renown clinical pathogens. This limits the recommendations for most anaerobic bacteria (Table S6). The FC/MIC ratio is more convenient for the estimation of the potential efficacy of poorly absorbed drugs. This ratio is particularly relevant in liver diseases associated with endogenous ethanol production, where gut microbiota plays a crucial role.

According to our results, amphotericin B was the most suitable for yeast. Furthermore, it has been shown to be highly effective in *in vitro* MAFLD models (28,29). Finally, its fecal concentration for the treatment of digestive candidiasis (2 g/day) was estimated at 60 μg/g of stool (24), which gives a ratio of the fecal concentration to the MIC of at least greater than 1 log_10_ (Figure 2) (15 to 30-fold (Table S6)).

**Figure 2.**
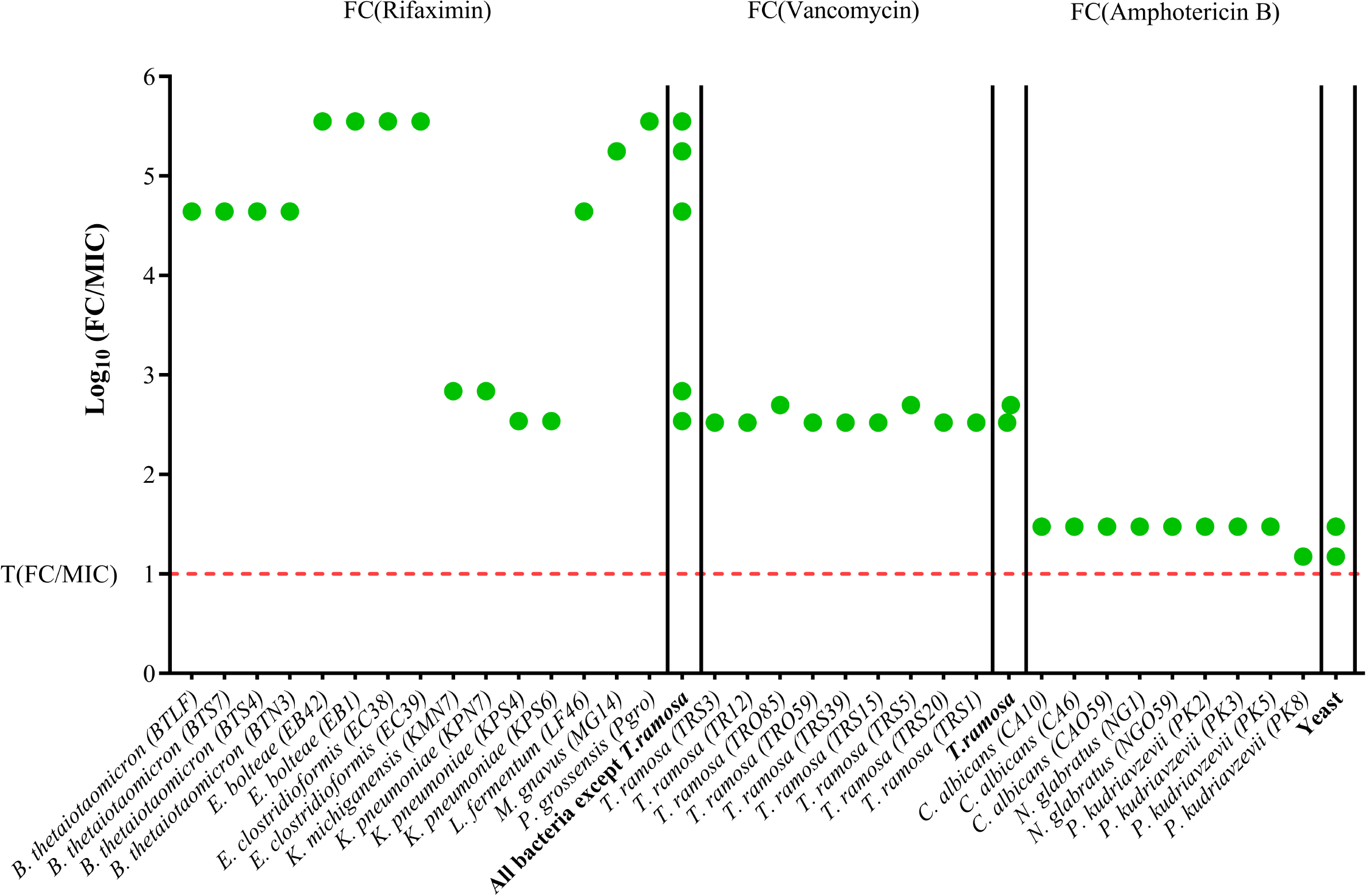
Potential *in vivo* efficacy of rifaximin, vancomycin and amphotericin B. Ratio of the fecal concentration after the recommended dose of amphotericin B, rifaximin and vancomycin to the MIC obtained after antifungal and antibiotic susceptibility tests. FC: Fecal concentration, MIC: Minimal inhibitory concentration, T: Threshold.

For most bacteria, rifaximin appeared as the better choice. Rifaximin is a water-soluble antibiotic that is poorly absorbed through the gut barrier and has broad-spectrum bactericidal activity (30). The fecal concentration of rifaximin needed to prevent relapses in encephalopathy (1100 mg/day) is estimated to be 11000 μg/g of stool (25). Therefore, the fecal concentration/MIC (FC/MIC) ratio ranged from 2 to 5 log_10_ (347 to 355,000-fold greater than the rifaximin FC/MIC (Table S6)). This ratio was highly variable depending on each genus and species: 2–3 log_10_ for *K. pneumoniae*; 4–5 log_10_ for *B. thetaiotaomicron* and *L. fermentum;* and > 5 log_10_ for all *Enterocloster* strains, *M. gnavus* and *P. grossensis* (Figure 2).

However, this ratio could not be estimated for *T. ramosa,* as its MIC was >256 µg/mL. Therefore, vancomycin seemed to be the most suitable antibiotic for *T. ramosa*. Furthermore, vancomycin could be used with the recommended posology for *Clostridium difficile* infection (CDI), as *T. ramosa* was previously known as *Clostridium ramosum* prior to reclassification. The fecal concentration of vancomycin used to treat CDI (500 mg/day) has been estimated at 1000 μg/g of stool (26), which resulted in a FC/MIC ratio > 2 log_10_ (333–500-fold higher than *T. ramosa* MICs (Table S6)) for all 9 *T. ramosa* strains (Figure 2). Notably, all these ratios were at least 15-fold higher than the MICs obtained (Table S6).

### Vancomycin resistant gene in *E. clostridioformis* align with phenotypic profile

The assessment of resistance genes towards rifaximin and vancomycin is crucial before the implementation of antibiotic therapy. Using AMRFinder Plus, which detects all resistance genes present in the NCBI database with at least 90% identity and contain gene *arr* alleles, we did not identify any *arr* genes in the bacterial genomes examined (Tables S7). *T. ramosa* strains were the only ones with high resistance to rifampicin and rifaximin (> 256 µg/mL, Table S5). Due to the lack of susceptible *T. ramosa* strain and the important phylogenetical distance between *T. ramosa* and main species studied in this context (*M. tuberculosis* or *E. coli* or *S. aureus*) (31), the mapping of rpoB regions was performed with *T. ramosa* genome of reference (ASM1672878v1). This mapping showed high similarities between our genomes and the reference one (only two mutations, Table S8) with no mutations found in the RRDR region (Table S8).

However, vancomycin resistance genes were detected in the genomes of *E. clostridioformis*. Specifically, strains EC38 and EC39 possessed four and ten vancomycin resistance genes, respectively (Table S7). These results are consistent with the *in vitro* vancomycin susceptibility profile observed for *E. clostridioformis* EC39 (Table S5). Resistance genes against macrolides, streptogramin, lincosamide and tetracycline were the most important genes within the studied genomes (Table S7).

## Discussion

The increasing incidence of liver diseases and the instrumental role of gut dysbiosis in their pathogenesis highlight the need for improved therapeutic approaches. Therefore, in this study, we characterized species enriched in these pathologies and assessed their ethanol production and tolerance as well as antimicrobial susceptibility. We found that gut microbial species enriched in the intestinal microbiota of patients with liver diseases produce significant quantities of ethanol *in vitro*. Notably, most of them are capable of both producing ethanol and demonstrating ethanol tolerance, as reported by Yuan et al. (10). Previous studies have reported ethanol production by yeasts (14), *K. pneumoniae* (10), *E. bolteae* (16), *L. fermentum* and *M. gnavus* (15), as well as ethanol tolerance in yeast and *L. fermentum* at concentrations exceeding 10% (32,33). This finding indicates a potentially vicious cycle in diseases such as ABS and MASH when a high-carbohydrate diet is sustained (Figure 3). To our knowledge, this study is the first to report *in vitro* ethanol production by *E. clostridioformis*, *K. michiganensis, P. grossensis*, and *T. ramosa*. Additionally, this is the first report of *E. clostridioformis* growth in the presence of 10% ethanol.

**Figure 3.**
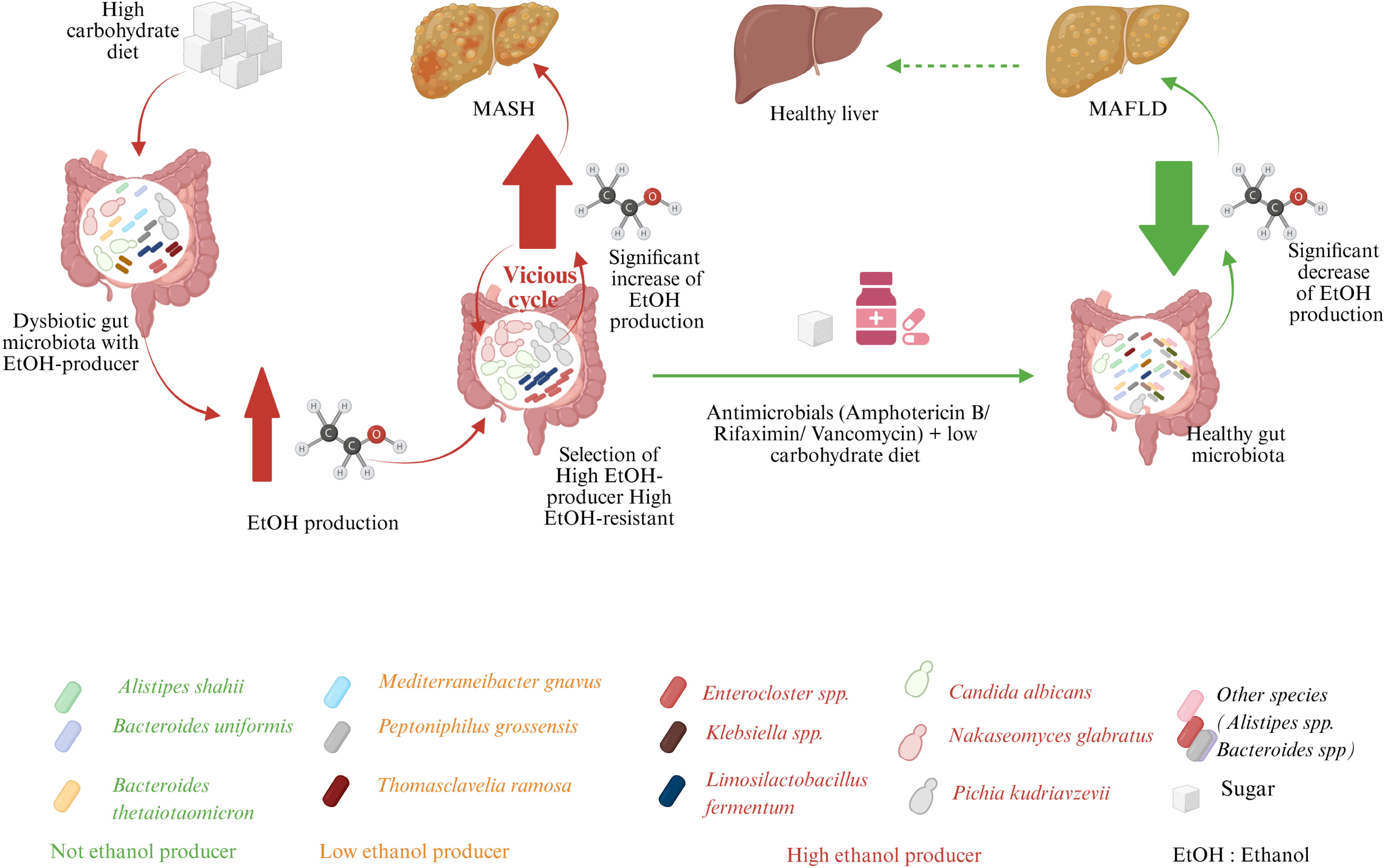
A vicious cycle caused by ethanol-producing ethanol-tolerant species due to a maintained high-glucose diet and the potential utilization of low-carbohydrate diet combined with antimicrobials as gut microbiota-targeted and precision medicine to disrupt the cycle.

Although anaerobic bacteria constitute the predominant microbial community within the gut microbiota (34), previous studies have used culture conditions that are not suitable for the isolation of strictly anaerobic bacteria such as *Enterocloster* that are enriched in the microbiota of patients with liver diseases. For instance, Yuan *et al.* (10) used yeast peptone dextrose (YPD) medium with a 24-hour incubation under both aerobic and anaerobic conditions, which did not allow the isolation of strictly anaerobic bacteria. This underscores the importance of using broader culture methods, such as culturomics, to better identify potential pathobionts in diseases such as MASH.

According to our findings, ethanol production varies with the species and glucose concentration in the growth medium. For instance, while *K. pneumoniae* increased ethanol production as the glucose concentration increased, *Enterocloster* decreased ethanol production under the same conditions. These results suggest the influence of glucose intake on endogenous ethanol production and suggest that carbohydrate intake may play a key role in the management of these diseases (35).

Enriched species in the intestinal microbiota of MASH patients, including yeast, *Enterocloster*, *Klebsiella* and *T. ramosa,* produce ethanol and hence may contribute to MASH pathogenesis. Xue et al. previously reported that reducing the abundance of ethanol-producing bacteria significantly using antibiotics could be a novel therapeutic approach (12). In our study, these pathobionts demonstrated high susceptibility to nonabsorbable antibiotics such as amphotericin B, rifaximin, and vancomycin (FC/MIC = 15–355,000), all of which have established efficacy in the treatment of various conditions (29, 36, 37). Nonetheless, it is noteworthy that fecal antimicrobial concentrations may vary because of individual patient characteristics and comorbidities (26).

Most importantly, in *in vivo* models for the treatment of MASH, amphotericin B, rifaximin and vancomycin have been demonstrated to reduce steatohepatitis and fibrosis (29, 36, 37). In clinical practice, patients diagnosed with both ABS and MASH have been successfully managed with antimicrobials in combination with low-carbohydrate diets and probiotics (Table 2). In particular, fluconazole is the most prescribed antifungal agent in these scenarios in this context (Table 2).

**Table 2.** Cases of ABS cured by antimicrobials in literature.

| Clinical disease(s) | Medical history | Max BAC (g/L) | Pathobiont(s) | Antimicrobial agents |  | Outcome | References |
| --- | --- | --- | --- | --- | --- | --- | --- |
| Antifungal |  |  |  |  |  |  |  |
| ABS | NA | 4.1 | *Saccharomyces cerevisiae*,<br>*Candida intermedia*,<br>*Klebsiella pneumoniae* and<br>*Enterococcus faecalis* | Micafungin | Cure | BAC (0 g/L) | Saverimuttu et al, 2019 (37) |
| ABS | NA | 4 | *Saccharomyces cerevisiae*,<br>*Candida albicans*, *Candida parapsilosis* | Micafungin | Cure | BAC (0 g/L) and no fungal growth | Malik et al, 2019 (38) |
| ABS | NA | 4 | *Saccharomyces cerevisiae* | Fluconazole then Nystatin | Cure | BAC (0 g/L) and negative stool cultures | Cordell et al, 2013 (39) |
| ABS | SGS | 3.5 | *Nakaseomyces glabratus* and<br>*Saccharomyces cerevisiae* | Fluconazole | Cure | Symptoms resolved | Dahshan et al, 2001 (40) |
| ABS | NA | 2.5 | *Candida albicans* and<br>*Candida krusei* | Trichomycin | Cure | Symptoms resolved | Kaji et al, 1984 (case 1) (41) |
| ABS | SBS | 0.7 | *Candida kefyr* and<br>*Saccharomyces cerevisiae* | Fluconazole | Cure | Symptoms resolved | Jansson-Nettelbladt et al, 2006 (42) |
| Antifungal and Antibacterial |  |  |  |  |  |  |  |
| ABS* | CIPO and SIBO | 1.1 | *Candida albicans* and<br>*Saccharomyces cerevisiae* | Fluconazole and Antibiotics | Cure | BAC (0 g/L) | Spinucci et al, 2006 (43) |
| Antibacterial |  |  |  |  |  |  |  |
| ABS | NA | 4 | *Klebsiella pneumoniae*,<br>*Klebsiella quasipneumoniae* and<br>*Klebsiella variicola* | Levofloxacin | Cure | BAC (0 g/L) | Xue et al, 2023 (12) |
| ABS* | NA | 4 | *Klebsiella pneumoniae* | Antibiotic | Cure | Recovery and MASH severity alleviated | Yuan et al, 2019 (10) |

ABS: Auto-brewery syndrome, Antibiotic: details not precise by authors, BAC: Blood alcohol concentration, CIPO: Chronic intestinal pseudo-obstruction, SBS: Short bowel syndrome, SGS: Short gut syndrome, SIBO: Small intestinal bacterial overgrowth, \*: respectively MAFLD and MASH

Antibiotic resistance represents a significant concern that must be addressed during antibiotic therapy. This underscores the importance of analyzing the resistome of isolated species of interest to minimize the emergence of resistance. Here, genomic resistome analysis further supported the potential application of rifaximin and vancomycin for managing MASH associated with bacterial ethanol producers based on the principal bacteria isolated.

The rpoB alignment of *T. ramosa* did not elucidate the nature of the resistance. Additionally, the lack of *T. ramosa* strain susceptible to rifampicin and or rifaximin evoked a natural resistance of these species. For a better understanding of resistance mechanisms identified here, further investigations should investigate and try to decipher vancomycin resistant genes in *E. clostridioformis* and the phenotypic resistance of *T. ramosa* to rifampicin and rifaximin.

Furthermore, this study highlights the importance of developing a microbiota-targeted and precision medicine approach for managing the gut microbiota dysbiosis associated with liver diseases (13). As previously discussed, compared with other species, certain species may produce more ethanol, exhibit greater tolerance to ethanol, or display increased resistance to antibiotics. This variation highlights the necessity for personalized, microbiota-targeted treatments. Ideally, one or two antimicrobials of interest can be used based on the patient’s microbial profile.

Given the potential for antibiotic resistance in the identified species and the possibility of significant changes in the composition of the microbiota due to antibiotic therapy (37), further research involving animal models and eventually human models is essential to validate the beneficial effects of combined antimicrobial agents in this context.

A limitation of this study is the relatively small number of strains analyzed for each species. A greater number of strains would strengthen the evidence for ethanol production and tolerance at the species level. Furthermore, an animal model would substantially enhance our understanding of how these species influence the development and progression of liver diseases, whether through individual or synergistic effects. However, besides all these limits, our work enables a full characterization of 33 strains associated with liver diseases as well as their disposition for the scientific community. These findings may offer valuable insights for managing hepatic disorders and could potentially have implications for liver cancer treatment.

## Conclusions

Most gut microbes associated with liver diseases in this study exhibited both tolerance to ethanol and the ability to produce it. The enrichment and characterization of endogenous ethanol-producing microbes, including *C. albicans*, *N. glabratus*, *P. kudriavzevii*, *E. clostridioformis*, *E. bolteae*, *L. fermentum*, *K. pneumoniae*, *K. michiganensis*, *T. ramosa* and *M. gnavus,* provide evidence that these species may play a significant role in liver diseases. Consequently, gut microbiota may serve as a target for precision medicine strategies tailored to individual patients. Potential antimicrobial therapies could include amphotericin B, rifaximin, and vancomycin. These findings may offer valuable insights for managing hepatic disorders and potentially have implications for cancer treatment.

## Data Availability

All the 33 genomes of the 33 bacterial strains of species enriched in the intestinal microbiota of patients with liver diseases are available in NBCI under BIOPROJECT: PRJNA1254750, except Q9705 and QA0666 which were previously deposited under the BIOPROJECT: PRJEB76822.
The authors confirm that the data supporting the findings of this study are available within the article and its supplementary materials.
All the 36 strains (33 associated with liver diseases and 3 associated with controls) reported in this study are publicly available and can be ordered in our microbial collection, the CSUR https://csur.eu/ and by mail.

## Abbreviations

ABS: Auto-brewery syndrome
COS: Columbia plus 5% sheep blood
EtOH: Ethanol
FC: Fecal concentration
HS-GC-MS: Headspace gas-gas chromatography coupled with mass spectrometry
MIC: Minimal inhibitory concentration
YPG: Yeast peptone glucose

## Data deposition

All the 33 genomes of the 33 bacterial strains of species enriched in the intestinal microbiota of patients with liver diseases are available in NBCI under BIOPROJECT: PRJNA1254750, except Q9705 and QA0666 which were previously deposited under the BIOPROJECT: PRJEB76822.

## Data availability

The authors confirm that the data supporting the findings of this study are available within the article and its supplementary materials.

## Strains availability

All the 36 strains (33 associated with liver diseases and 3 associated with controls) reported in this study are publicly available and can be ordered in our microbial collection, the CSUR https://csur.eu/ and by mail. The number of each strain is reported in Table 1.

## Acknowledgments

We thank Nicolas ORAIN for technical help as well as Hanh NGUYEN THI MY from the hepatology unit. We also thank Dr Fadi BITTAR for his help in antimicrobials resistance genes interpretations. Figure 3 (VN28ZL02DT) were Created in https://BioRender.com. Claude ai (version Haiku 4.7) and Copilot ai (version: bizchat.20251030.51.2) were used only for rephrasing. The final meaning was checked and ensured that the message that we went to vehiculate was intact.

## Author contributions

Writing – original draft: AIA, MB, CA, SB; Writing – review & editing: MM, MTA, JCL, GD, RG; Conceptualization: RG, MM; Investigation: AIA, BM,NA RMW, AH, JMR, SR, GD; Data curation: BM, RMW, AIA, MB, CA, SB; Methodology: MM, RG, MTA; Supervision: MM, RG, MTA; Formal analysis: NA, AIA; Project administration: RG, MM; Validation: RG, MM, MTA; Funding acquisition: RG, MM; Resources: PB, RG, LC; Visualization: AIA, MM, MTA.

