## Supplementary data for "Potential Therapeutic Options for Ethanol-Producing Ethanol-Resistant Gut Microbes associated with Liver Diseases"

### Supplementary Materials and Methods

#### Ethanol assay

The quantification of ethanol production by bacteria was carried out by inoculating 250 µL of 0.5 McFarland for aerobic bacteria and 1 McFarland for anaerobic bacteria in 5 mL of liquid Columbia with 5% sheep's blood (COS). For yeasts, 250 µL of 0.5 McFarland suspension was inoculated into 5 mL of and incubated for 24 hours at 30°C.

COS broth is composed of : 5 g/L glucose (MP, Ilkirch, France), 12 g/L tryptone (Sigma-Aldrich, St Louis, USA), 5 g/L protease peptone (Sigma-Aldrich, St Louis, USA), 3 g/L yeast extract (bioMérieux, Marcy l'Etoile, France), 3 g/L beef extract (Sigma-Aldrich, St Louis, USA), 1g/L potato starch (Sigma-Aldrich, St Louis, USA), 5 g/L sodium chloride (Sigma-Aldrich, St Louis, USA), adjusted to pH 6.5 and 5% defibrinated horse blood (Oxoid). Our anaerobic bacteria tubes were degassed for 3 minutes with a gas mixture (H2/N2/CO2) prior to inoculation. Inoculums were incubated at 37°C for 24 hours for aerobic bacteria and 48 hours for anaerobic bacteria.

Assays were performed in triplicate for each strain, together with a negative control consisting of 5 mL of inoculated medium with 250 µL of 0.85% NaCL solution (bioMérieux, Marcy l'Etoile, France). After incubation, 1 mL of each culture and negative control were poured into 20 mL Headspace glass vials.

Twelve ethanol solutions with concentrations ranging from 0.25 to 100 mM, together with a blank made up entirely of water, formed our calibration range. To each solution in our range, as well as to each inoculum, 50 µL of 5 mM 2-isopropanol, our internal standard, was added.

Dosing of the various alcohols (methanol, ethanol, isopropanol and propanol) was carried out using a Headspace gas chromatography-mass spectrometry (HS-GC/MS) system (Perkin Elmer, Villebon sur Yvette, France) in a Swafer D7 set-up, combining an HS110 headspace injector, a Clarus 690 gas chromatograph and an SQ8T mass spectrometer. After dosage, each inoculum was re-inoculated on COS agar to ensure that there was no contamination and re-incubated according to the previously defined parameters.

**Ethanol tolerance**

To evaluate ethanol tolerance, we added 250 µL of 1 McFarland suspension to 5 mL of liquid COS containing 0%, 5% and 10% pure ethanol after 3 min degassing for anaerobic bacteria and 0.5 McFarland for aerobic bacteria. For yeast, 250 µL of 0.5 McFarland yeast suspension was added in liquid Sabouraud (Oxoid) with 0%, 5% and 10% pure ethanol.

After incubation at 37°C for 24 hours for aerobic bacteria, 48H for anaerobic bacteria, 100 µL of each bacterial inoculum was inoculated on COS agar (bioMérieux, Marcy l'Etoile, France) and then re-incubated under the same conditions as before with gas packs (Becton Dickinson, Sparks, USA) added for anaerobic bacteria.

For yeasts, after 24 hours at 30°C, 100 µL of each inoculum was inoculated on Sabouraud agar supplemented with gentamycin and chloramphenicol (bioMérieux, Marcy l'Etoile, France), then re-incubated under the same conditions as above.

**Antibiogram by ETEST**

Twenty-five antibiotics including: amikacin, amoxicillin, ceftazidime, ceftriaxone, ciprofloxacin, clindamycin, colistin, daptomycin, doxycycline, ertapenem, fosfomycin, gentamycin, imipenem, levofloxacin, linezolid, metronidazole, nitrofurantoin, norfloxacin, oxacillin, benzylpenicillin, rifampicin, teicoplanin, tobramycin, trimethoprim-sulfamethoxazole and vancomycin were tested by ETEST (bioMérieux).

For aerobic bacteria, a suspension of 0.5 McFarland in 2 mL 0.85% NaCl solution (bioMérieux) was spread on COS agar (bioMérieux) with a sterile swab, while a suspension of 1 McFarland in 2 mL 0.85% NaCl solution (bioMérieux) was used for anaerobic bacteria. The test was performed in duplicate using 2 antibiotic strips for each strain, and positive control without antibiotics was also performed. Anaerobic bacteria were placed in zip bags (Becton Dickinson, Sparks, USA) containing gas packs (Becton Dickinson) to mimic the anaerobic condition. Minimum inhibitory concentration (MIC) values were read after 20 hours incubation at 37°C for aerobic bacteria and 48 hours at 37°C for anaerobic bacteria.

**Rifaximin MIC (Agar dilution)**

Mueller Hinton agar medium composed of: 2 g/L meat extract (Sigma-Aldrich, St Louis, USA), 17.5 g/L casein acid hydrolase (Becton Dickinson, Le Pont de Claix, France), 1.5 g/L starch (Sigma-Aldrich, St Louis, USA) and 15 g/L agar (Sigma-Aldrich, St Louis, USA) was prepared for aerobic bacteria.

Brucella agar medium consisted of: 23 g/L Brucella Agar (Sigma-Aldrich, St Louis, USA) supplemented with hemin 5 µg/mL (Sigma-Aldrich), vitamin K1 1 µg/mL (Sigma-Aldrich) and 5% defibrinated and thawed horse blood (Oxoid), was prepared for testing anaerobic bacteria.

Starting with a stock solution of rifaximin at 2560 µg/mL, a cascade dilution to one-half the concentration of 0.3 µg/mL was performed in culture broth. A second one-tenth dilution of each rifaximin solution (256 µg/mL to 0.03 µg/mL) was made in autoclaved agar media with the addition of 5% defibrinated and thawed horse blood (Oxoid) for anaerobic bacteria.

They were individually poured into annotated 90 mm petri dishes divided into 23 dials under microbiological station type 2 (PSM II). Rifaximin-free agar media constituted our positive controls, and an uninoculated agar was used as our negative control.

2 µL of 0.5 McFarland suspension in a 2 mL solution of 0.85% NaCl (bioMérieux) of each strain was spotted on the corresponding quadrant on the agar plates. The test was performed in duplicate. The strains were incubated following the same procedure as above.

**Antifungal**

*VITEK 2*

For each strain, a suspension of fresh colonies between 1.80 and 2.20 in 3 mL of 0.45% NaCl saline (bioMérieux, Marcy L'Etoile, France) was made.

*Fluconazole ETEST*

The fluconazole susceptibility test was carried out by ETEST (bioMérieux). A suspension of 0.5 McFarland of fresh colony was made in a 0.85% NaCl solution (bioMérieux). For each strain, the suspension was streaked on Sabouraud agar supplemented with gentamycin and chloramphenicol (bioMérieux), followed by two strips impregnated with fluconazole. Agar plates were incubated at 30°C for 24 hours before the results were read.

**Potential efficacy of amphotericin B, rifaximin and vancomycin in vivo**

To assess the potential efficacy of amphotericin B on yeast, its fecal concentration after a digestive candidiasis (2 g/day) has been estimated at 60 µg/g of stool. The fecal concentration of rifaximin in humans for a treatment of 800 mg/day is 7961 µg /g after the first day of treatment. In France, Tixtar (1100 mg/day) is the commercial form of rifaximin used for the prevention of relapses of hepatic encephalopathy. Taking this dosage into account, we had estimated the concentration of rifaximin in stools at 11000 µg/g. The fecal concentration of vancomycin (1000 µg/g) during treatment of *C. difficile* infection with vancomycin (500 mg/day) was used.

**Bacterial genomes sequencing**

Genomic DNA was sequenced using MiSeq (Illumina, San Diego, CA) with paired end reads and barcoding for multiplexing. Each genome was diluted to 1 ng, fragmented and tagged via tagmentation, then amplified by 12-cycle PCR to add dual-index barcodes. Libraries were purified with AMPure XP beads (Beckman Coulter Inc, Fullerton, CA, USA), normalized per Nextera XT protocol (Illumina), pooled, and sequenced on MiSeq (Illumina) in a single 39-hour run using 2x250-bp reads (MiSeq Reagent Kit V2-500 cycles).

**Bacterial genomes assembling**

Direct quality control was performed on the sequence reads generated by MiSeq (Illumina), utilizing FastQC to visualize sequence quality (1). Trimmomatic v0.39 (2) was used to trim and enhance read quality. Trimmed reads were assembled with Unicycler v0.4.8 (3), and scaffolds shorter than 800 bp were excluded. Scaffolds with a depth lower than 25% of the average were identified as potential contaminants and removed.

**Supplementary Figures**

**Figure S1. Comparison of ethanol production species enriched in cases and species enriched in controls**

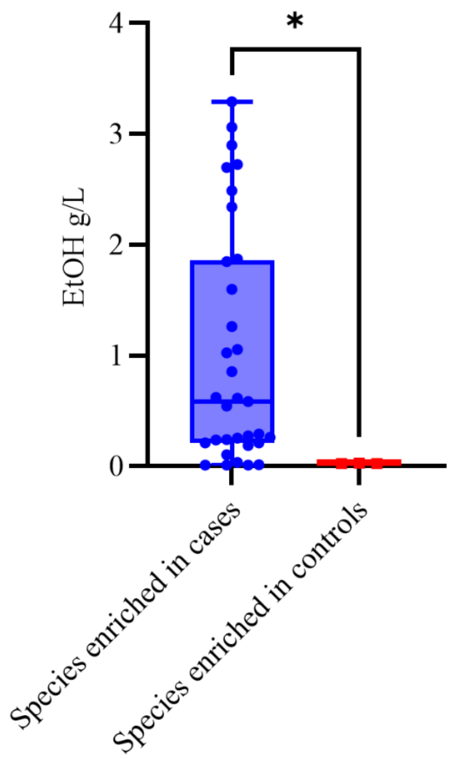

Fig. S1. Dosage was performed via gas chromatography coupled with mass spectrometry. Level of significance *: p = 0.0286 (two-tailed Mann Whitney test). Abbreviation: EtOH: Ethanol produced dose.

**Figure S2. Comparison of ethanol production between yeast and bacteria**

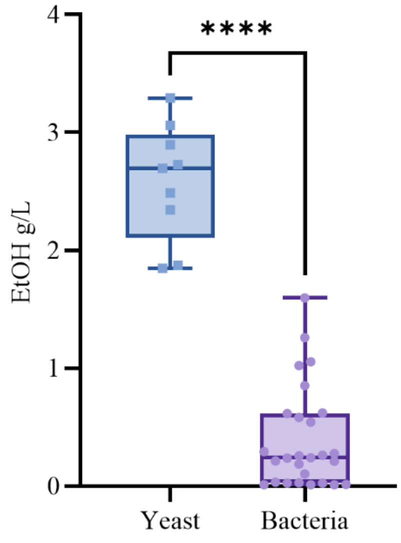

Fig. S2. Dosage was performed via gas chromatography coupled with mass spectrometry. Level of significance: ****: $p<0.0001$(two-tailed Mann Whitney test). Abbreviation: EtOH: Ethanol produced dose.

**Figure S3**. **Ethanol produced in 20g/L of glucose by yeast and bacteria**

A. B.

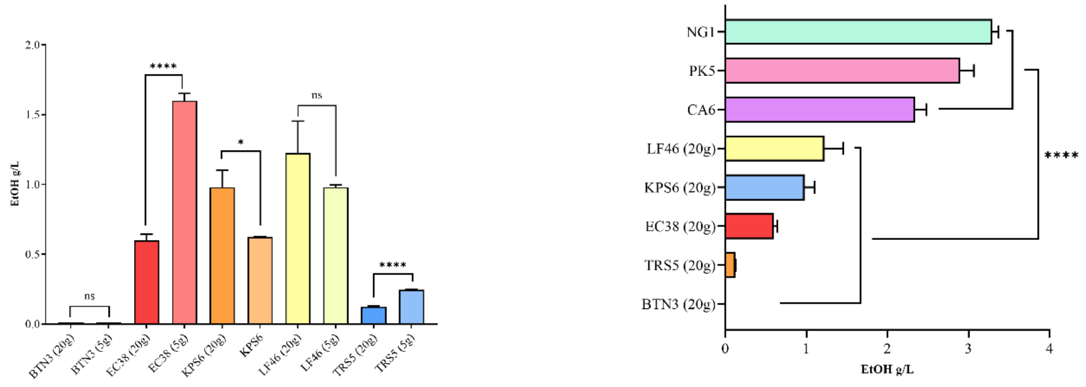

**Fig. S3.** Dosage was performed by gas chromatography coupled with mass spectrometry. (A) Comparison of ethanol production of ethanol in 5 g/L vs 20 g/L of glucose by bacteria (EtOH: Ethanol produced dosed). Level significance: ****: p<0.0001 (Unpaired t test with Welch's correction, two-tailed); *: p=0.0359 (Unpaired t test with Welch's correction, two-tailed); ns: p=0.2070 (Unpaired t test with Welch's correction, two-tailed). (B) Comparison of ethanol production by yeast and bacteria in 20 g/L (EtOH: Ethanol produced dosed). Level significance: ****: p<0.0001 (two-tailed Mann Whitney test).

### Supplementary Tables

#### Table S1. Mean ethanol quantity and standard deviation produced by all strains

| Strains | Mean (g/L) | Standard deviation |
|---|---|---|
| *Alistipes shahii T1 (AST1)* | 0.024 | 0.00046 |
| *Alistipes shahii S4 (ASS4)* | 0.028 | 0.00184 |
| *Bacteroides uniformis S4 (BUS4)* | 0.025 | 0.00096 |
| *Bacteroides thetaiotaomicron LF 115 (BTLF)* | 0.012 | 0.00027 |
| *Bacteroides thetaiotaomicron S7 (BTS7)* | 0.014 | 0.00070 |
| *Bacteroides thetaiotaomicron S4 (BTS4)* | 0,013 | 0.0017 |
| *Bacteroides thetaiotaomicron N3 (BTN3)* | 0.013 | 0.0013 |
| *Enterocloster bolteae 1' S8 (EB1)* | 1.26 | 0.10 |
| *Enterocloster bolteae 42 S1R (EB42)* | 1.05 | 0.064 |
| *Enterocloster clostridioformis S8 39 (EC39)* | 0.85 | 0.15 |
| *Enterocloster clostridioformis 38' S1R (EC38)* | 1.60 | 0.053 |
| *Klebsiella michiganensis N7 (KMN7)* | 0.54 | 0.023 |
| *Klebsiella pneumoniae N7 (KPN7)* | 0.58 | 0.022 |
| *Klebsiella pneumoniae S4 (KPS4)* | 0.62 | 0.0087 |
| *Klebsiella pneumoniae S6 (KPS6)* | 0.62 | 0.0043 |
| *Limosilactobacillus fermentum 46 S8-S9R (LF46)* | 1.02 | 0.014 |
| *Mediterraneibacter gnavus 14 S8 R (MG14)* | 0.10 | 0.0032 |
| *Peptoniphilus grossensis 58' S1R (Pgro)* | 0.034 | 0.0012 |
| *Thomasclavelia ramosa S3R (TRS3)* | 0.28 | 0.0053 |
| *Thomasclavelia ramosa 12' S8 (TR12)* | 0.21 | 0.018 |
| *Thomasclavelia ramosa O85 LT (TRO85)* | 0.24 | 0.0086 |
| *Thomasclavelia ramosa O59 (TRO59)* | 0.26 | 0.025 |
| *Thomasclavelia ramosa S5R (TRS5)* | 0.26 | 0.0044 |
| *Thomasclavelia ramosa S20R (TRS20)* | 0.29 | 0.014 |
| *Thomasclavelia ramosa S39R (TRS39)* | 0.24 | 0.0073 |
| *Thomasclavelia ramosa S15R (TRS15)* | 0.21 | 0.013 |
| *Thomasclavelia ramosa S1R (TRS1)* | 0.19 | 0.016 |
| *Candida albicans N10 (CA10)* | 1.87 | 0.041 |
| *Candida albicans N6 (CA6)* | 2.34 | 0.14 |
| *Candida albicans O59 HF (CA059)* | 1.84 | 0.16 |
| *Nakaseomyces glabratus N1 (NG1)* | 3.29 | 0.077 |
| *Nakaseomyces glabratus O59HF (NGO59)* | 3.06 | 0.27 |
| *Pichia kudriavzevii N2 (PKN2)* | 2.70 | 0.10 |
| *Pichia kudriavzevii N3 (PKN3)* | 2.73 | 0.15 |
| *Pichia kudriavzevii N5 (PKN5)* | 2.90 | 0.17 |
| *Pichia kudriavzevii Nash8 (PKN8)* | 2.49 | 0.25 |

Ethanol dosage was performed by gas chromatography coupled with mass spectrometry

**Table S2. Ethanol concentration dosed in negative controls (only medium)**

| Medium | mM | g/L |
|---|---|---|
| Aerobes negative control | Not detected | Not detected |
| | Not detected | Not detected |
| | Not detected | Not detected |
| Anaerobes negative control | 0,10 (<LQ) | 0,0046 (<LQ) |
| | 0,11 (<LQ) | 0,00506 (<LQ) |
| | 0,09 (<LQ) | 0,00414 (<LQ) |
| Yeasts negative control | 0,007 (<LQ) | 0,000322 (<LQ) |
| | 0,003 (<LQ) | 0,000138 (<LQ) |
| | 0,003 (<LQ) | 0,000138 (<LQ) |

<LQ: alcohol was detected below limit of quantification

### Table S3. Growth in the presence of 0%, 5% and 10% ethanol

| | Genus/Species | Strains | 0% | 5% | 10% |
|---|---|---|---|---|---|
| Strains belonging to species enriched in controls | *Alistipes shahii* | *Alistipes shahii T1 (AST1)* | 1 | 1 | 0 |
| | | *Alistipes shahii S4 (ASS4)* | 1 | 0 | 0 |
| | *Bacteroides uniformis* | *Bacteroides uniformis S4 (BUS4)* | 1 | 1 | 0 |
| Strains belonging to species enriched in liver diseases | *Bacteroides thetaiotaomicron* | *Bacteroides thetaiotaomicron LF 115 (BTLF)* | 1 | 1 | 0 |
| | | *Bacteroides thetaiotaomicron S7 (BTS7)* | 1 | 1 | 0 |
| | | *Bacteroides thetaiotaomicron S4 (BTS4)* | 1 | 1 | 0 |
| | | *Bacteroides thetaiotaomicron N3 (BTN3)* | 1 | 1 | 0 |
| | *Candida albicans* | *Candida albicans N10 (CA10)* | 1 | 1 | 1 |
| | | *Candida albicans N6 (CA6)* | 1 | 1 | 1 |
| | | *Candida albicans O59 HF (CA059)* | 1 | 1 | 1 |
| | *Enterocloster spp.* | *Enterocloster bolteae 1' S8 (EB1)* | 1 | 1 | 0 |
| | | *Enterocloster bolteae 42 S1R (EB42)* | 1 | 0 | 0 |
| | | *Enterocloster clostridioformis S8 39 (EC39)* | 1 | 1 | 1 |
| | | *Enterocloster clostridioformis 38' S1R (EC38)* | 1 | 1 | 0 |
| | *Klebsiella spp.* | *Klebsiella michiganensis N7 (KMN7)* | 1 | 1 | 0 |
| | | *Klebsiella pneumoniae N7 (KPN7)* | 1 | 1 | 0 |
| | | *Klebsiella pneumoniae S4 (KPS4)* | 1 | 1 | 0 |
| | | *Klebsiella pneumoniae S6 (KPS6)* | 1 | 1 | 0 |
| | *Limosilactobacillus fermentum* | *Limosilactobacillus fermentum 46 S8-S9R (LF46)* | 1 | 1 | 1 |
| | *Mediterraneibacter gnavus* | *Mediterraneibacter gnavus 14 S8 R (MG14)* | 1 | 1 | 0 |
| | *Peptoniphilus grossensis* | *Peptoniphilus grossensis 58' S1R (Pgro)* | 1 | 1 | 0 |
| | *Pichia kudriavzevii* | *Pichia kudriavzevii N2 (PKN2)* | 1 | 1 | 1 |
| | | *Pichia kudriavzevii N3 (PKN3)* | 1 | 1 | 1 |
| | | *Pichia kudriavzevii N5 (PKN5)* | 1 | 1 | 1 |
| | | *Pichia kudriavzevii Nash8 (PKN8)* | 1 | 1 | 1 |
| | *Nakaseomyces glabratus* | *Nakaseomyces glabratus N1 (NG1)* | 1 | 1 | 1 |
| | | *Nakaseomyces glabratus O59HF (NGO59)* | 1 | 1 | 1 |
| | *Thomasclavelia ramosa* | *Thomasclavelia ramosa S3R (TRS3)* | 1 | 1 | 0 |
| | | *Thomasclavelia ramosa 12' S8 (TR12)* | 1 | 1 | 0 |
| | | *Thomasclavelia ramosa O85 LT (TRO85)* | 1 | 1 | 0 |
| | | *Thomasclavelia ramosa O59 (TRO59)* | 1 | 1 | 0 |
| | | *Thomasclavelia ramosa S5R (TRS5)* | 1 | 1 | 0 |
| | | *Thomasclavelia ramosa S20R (TRS20)* | 1 | 1 | 0 |
| | | *Thomasclavelia ramosa S39R (TRS39)* | 1 | 1 | 0 |
| | | *Thomasclavelia ramosa S15R (TRS15)* | 1 | 1 | 0 |
| | | *Thomasclavelia ramosa S1R (TRS1)* | 1 | 1 | 0 |

0: no colony grew, 1: at least one colony grew

**Table S4. Anti-fungal tests results**

| Antifungals | *Candida albicans* n=3 | *Nakaseomyces glabratus* n=2 | *Pichia kudriavzevii* n=4 |
|---|---|---|---|
| Fluconazole [a] | 1 | 8 | 24 - 32 |
| Voriconazole | ≤ 0.12 | ≤ 0.12 | ≤ 0.12 |
| Caspofungin | ≤ 0.12 - 0.25 | 0.25 | 0.5 |
| Micafungin | ≤ 0.06 | ≤ 0.06 | 0.12 |
| Amphotericin B | 0.5 | 0.5 | 0.5 - 4 |
| Flucytosin | ≤ 1 | ≤ 1 | 16 |

[a]: test done by Etest, n: number of strains, minimal inhibitory concentration (MIC) given in µg/mL

### Table S5. Antibiotic susceptibility tests results

| Antibiotics | | Anaerobic bacteria | | | | | | Aerobic bacteria |
|---|---|---|---|---|---|---|---|---|
| | | *B. thetaitaomicron* (n=4) | *Enterocloster* spp. (n=4)[a] | *L. fermentum* (n=1) | *M. gnavus* (n=1) | *P. grossensis* (n=1) | *T. ramosa* (n=9) | *Klebsiella* spp. (n=4)[b] |
| Aminoglycosides | Amikacin | >256 | 96 - >256 | 6 | >256 | 96 | >256 | 3 - 4 |
| | Gentamycin | >256 | <0.016 - 12 | 2 | 32 | 0.38 | 24 - 64 | 0.5 - 1 |
| | Tobramycin | >256 | 1.5 - >256 | 6 | >256 | 12 | 64 - 256 | 1.5 - 2 |
| Ansamycins | Rifampicin | 0.032 - 0.125 | <0.002 - 0.006 | 0.25 | <0.002 | <0.002 | >32 | >32 |
| | Rifaximin[c] | 0.25 | 0.031 | 0.25 | 0.062 | 0.031 | >256 | 16 - 32 |
| Carbapenems | Ertapenem | 0.19 - 0.50 | 0.032 - 0.19 | 0.125 | 1 | <0.002 | 0.25 - 0.50 | 0.003 |
| | Imipenem | 0.25 - 0.38 | 0.38 - 2 | 0.023 | 0.50 | 0.006 | 0.125 - 0.25 | 0.125 - 0.19 |
| Cephalosporins | Ceftazidim | 64 - >256 | 0.125 - >256 | 4 | 32 | 0.094 | 1 -2 | 0.094 - 0.125 |
| | Ceftriaxon | >256 | 0.25 - >256 | 8 | 4 | 0.064 | 0.094 - 0.25 | 0.023 - 0.064 |
| Fluoroquinolones | Ciprofloxacin | >32 | >32 | 2 | >256 | 3 | 6 - >32 | 0.023 - 0.032 |
| | Levofloxacin | >32 | >256 | 2 | >32 | 8 | 8 - >32 | 0.064 |
| | Norfloxacin | >256 | >256 | 24 | >256 | 6 | >256 | 0.19 - 0.25 |
| Glycopeptides | Teicoplanin | 12 -24 | 1.5 | >256 | 0.047 | <0.016 | 0.094 - 0.25 | >256 |
| | Vancomycin | 24 - 32 | 0.094 - 32 | 0.75 | 0.38 | 0.064 | 2 -3 | >256 |
| Lincosamides | Clindamycin | 0.094 - >256 | <0.016 - 0.38 | <0.016 | 0.125 | >256 | 0.19 - >256 | >256 |
| Miscellaneous agents | Colistin | >256 | >256 | 32 | >256 | >256 | >256 | 0.125 |
| | Daptomycin | >256 | 0.023 - 2 | 0.064 | 0.25 | 0.016 | 12 -32 | >256 |
| | Fosfomycin | >1024 | 6 - >512 | >1024 | >1024 | 2 | 4 - 16 | 16 - 48 |
| | Metronidazole | 0.125 - 0.38 | <0.016 | >256 | 0.094 | 0.047 | 0.19 - 8 | >256 |
| | Nitrofurantoin | 1.5 - 3 | 0.19 - 0.50 | 3 | 0.25 | 0.38 | 0.25 -1 | 16 - 48 |
| | Trimethoprim-sulfamethoxazole | 0.125 - >32 | 0.25 - >32 | >32 | >32 | >32 | 0.064 - >32 | 6 - >32 |
| Penicillins | Amoxicillin | 24 - >256 | 0.125 - 64 | 0.19 | 0.25 | <0.016 | 0.032 - 0.094 | 64 - 128 |
| | Benzylpenicillin | 16 - 256 | 0.25 - >256 | 0.125 | 0.38 | 0.032 | 0.047 - 0.50 | 48 - 96 |
| | Oxacillin | >256 | 6 - >256 | 3 | 8 | 0.19 | 1 - >256 | >256 |
| Oxazolidinones | Linezolid | 1 - 4 | 0.25 - 1 | 1 | 1 | 0.19 | 0.75 - 12 | >256 |
| Tetracyclines | Doxycyclin | 0.094 - 8 | 4 - 12 | 4 | 3 | 8 | 1.5 - 0.094 | 2 |

[a]: *Enterocloster bolteae* (n=2) and *Enterocloster clostridioformis* (n=2), [b]: *Klebsiella pneumoniae* (n=3) and *Klebsiella michiganensis* (n=1), [c]: Rifaximin susceptibility test was done by Agar dilution, minimal inhibitory concentration (MIC) is given in µg/mL

### Table S6. Potential efficacity of amphotericin B, rifaximin and vancomycin *in vivo* gut microbiota

| Species | Anaerobic bacteria | | | | | | | | | | | | Aerobic bacteria | | Yeasts | | |
|---|---|---|---|---|---|---|---|---|---|---|---|---|---|---|---|---|---|
| | *B. thetaitaomicron* | | *Enterocloster* spp. | | *L. fermentum* | | *M. gnavus* | | *P. grossensis* | | *T. ramosa* | | *Klebsiella* spp. | | *C. albicans* | *N. glabratus* | *P. kudriavzevii* |
| Poorly absorbed antimicrobials | RFX | VAN | RFX | VAN | RFX | VAN | RFX | VAN | RFX | VAN | RFX | VAN | RFX | VAN | AmB | AmB | AmB |
| MIC (µg/mL) | 0.25 | 24 - 32 | 0.031 | 0.094 - 32 | 0.25 | 0.75 | 0.062 | 0.38 | 0.031 | 0.064 | >256 | 2 -3 | 16 - 32 | >256 | 0.5 | 0.5 | 0.5 - 4 |
| FC (µg/g) | 11000 | 1000 | 11000 | 1000 | 11000 | 1000 | 11000 | 1000 | 11000 | 1000 | 11000 | 1000 | 11000 | 1000 | 60 | 60 | 60 |
| FC/MIC | 44000 | 31- 42 | 355000 | 10638 - 31 | 44000 | 1333 | 177000 | 2632 | 355000 | 15625 | NA | 333 - 500 | 347 - 688 | NA | 30 | 30 | 15- 30 |
| Interpretation in blood* | NA | NA | NA | NA | NA | NA | NA | NA | NA | NA | NA | NA | NA | NA | S = 100% | S = 100% | S = 50% |
| Interpretation in feces$^{\mu}$ | S = 100% | S = 100% | S = 100% | S = 100% | S = 100% | S = 100% | S = 100% | S = 100% | S = 100% | S = 100% | NA | S = 100% | S = 100% | NA | S = 100% | S = 100% | S = 100% |

AmB: Amphotericin B, RFX: Rifaximin; VAN: Vancomycin, S: percentage of susceptibility, MIC: Minimal inhibitory concentration, FC: Fecal concentration, NA: not available, *: based on EUCAST 2025 recommendations (4,5), $^{\mu}$: based on fecal concentrations (6–8)

**Table S7. Antibiotic resistance gene detected *in silico* with AMRFinderPlus**

| Strains | Macrolide/ Streptogramin/ Lincosamide | ESBL/ Beta-lactam | Tetracycline | Vancomycin | Kanamycin | Phenicol/ Quinolone | Fosfomycin | Streptothricin | Aminoglycoside | Trimethoprim | Rifampicin |
|---|---|---|---|---|---|---|---|---|---|---|---|
| *Bacteroides thetaiotaomicron LF 115 (BTLF)* | erm(F) | | tet(X2)* | | | | | | | | |
| *Bacteroides thetaiotaomicron S7 (BTS7)* | erm(F),mef(En2)*, lnu(AN2) | cfxA4* | tet(Q)*, tet(X2)* | | | | | | | | |
| *Bacteroides thetaiotaomicron S4 (BTS4)* | erm(G)* | cfxA3 | tet(Q) | | | | | | | | |
| *Bacteroides thetaiotaomicron N3 (BTN3)* | | | | | | | | | | | |
| *Enterocloster bolteae 42 S1R (EB42)* | erm(B) | | tet(O)* | | | | | | | | |
| *Enterocloster bolteae 1' S8 R (EB1)* | | | tet(40)* | | | | | | | dfrF | |
| *Enterocloster clostridioformis 38' S1R (EC38)* | | | tet(32)* | vanR-D*,vanS-D*,vanX-D* | | | | | | | |
| *Enterocloster clostridioformis S8 39 (EC39)* | erm(B)* | | tet(W) | vanB,vanH-B*,vanR-B,vanR-D*,vanS-B*,vanS-D*,vanW-B,vanX-B,vanX-D*,vanY-B | | | | | | | |
| *Klebsiella michiganensis N7 (KMN7)* | | blaOXY-1-7 | | | aph(3')-Ia | oqxA*,oqxB9* | fosA7.3 | | | | |
| *Klebsiella pneumoniae N7 (KPN7)* | | blaSHV-1 | | | | oqxA10,oqxB19 | fosA | | | | |
| *Klebsiella pneumoniae S4 (KPS4)* | | blaSHV-28 | | | | oqxA,oqxB20* | fosA | | | | |
| *Klebsiella pneumoniae S6 (KPS6)* | | blaSHV-11 | | | | oqxA*,oqxB5 | fosA | | | | |
| *Limosilactobacillus fermentum 46 S8-S9R (LF46)* | | | | | | | | | | | |
| *Mediterraneibacter gnavus 14 S8 R (MG14)* | | | tet(O)* | | | | | | | | |
| *Peptoniphilus grossensis 58' S1R (Pgro)* | erm(A), Isa(C) | | tet(M) | | aph(3')-IIIa | | | sat4 | ant(6)-Ia | | |
| *Thomasclavelia ramosa S3R (TRS3)* | | | tet(44)* | | | | | | | | |
| *Thomasclavelia ramosa 12' S8 (TRS8)* | | | | | | | | | | | |
| *Thomasclavelia ramosa O85 LT (TRO85)* | | | | | | | | | | | |
| *Thomasclavelia ramosa O59 (TRO59)* | | | | | | | | | | | |
| *Thomasclavelia ramosa S5R (TRS5)* | | | | | | | | | | | |
| *Thomasclavelia ramosa S20R (TRS20)* | erm(B) | | | | | | | | aadE | dfrF | |
| *Thomasclavelia ramosa S39R (TRS39)* | | | tet(44)* | | | | | | | | |
| *Thomasclavelia ramosa S15R (TRS15)* | erm(B) | | | | | | | | | | |
| *Thomasclavelia ramosa S1R (TRS1)* | erm(B) | | | | | | | | aadE | dfrF | |

Resistance genes detected with at least 90% of identity. *: resistance genes detected with less than 100% identity

### **Table S8. Mutations found in *rpoB* gene (rifaximin and rifampicin resistance)**

| **Species** | **Mutation** |
|---|---|
| *Thomasclavelia ramosa* Type strain | Reference |
| *Thomasclavelia ramosa S3R (TRS3)* | *rpoB* sequence identical to the reference |
| *Thomasclavelia ramosa 12' S8 (TRS8)* | |
| *Thomasclavelia ramosa O85 LT (TRO85)* | |
| *Thomasclavelia ramosa S5R (TRS5)* | |
| *Thomasclavelia ramosa S20R (TRS20)* | |
| *Thomasclavelia ramosa S15R (TRS15)* | |
| *Thomasclavelia ramosa S1R (TRS1)* | |
| *Thomasclavelia ramosa O59 (TRO59)* | E206K |
| *Thomasclavelia ramosa S39R (TRS39)* | E213V |

E: Glutamate, K = Lysine, V = Valine
